# Using influenza surveillance networks to estimate state-specific case detection rates and forecast SARS-CoV-2 spread in the United States

**DOI:** 10.1101/2020.04.01.20050542

**Authors:** Justin D. Silverman, Nathaniel Hupert, Alex D. Washburne

## Abstract

Detection of SARS-CoV-2 infections to date has relied on RT-PCR testing. However, a failure to identify early cases imported to a country, bottlenecks in RT-PCR testing, and the existence of infections which are asymptomatic, sub-clinical, or with an alternative presentation than the standard cough and fever have resulted in an under-counting of the true prevalence of SARS-CoV-2. Here, we show how publicly available CDC influenza-like illness (ILI) outpatient surveillance data can be repurposed to estimate the detection rate of symptomatic SARS-CoV-2 infections. We find a surge of non-influenza ILI above the seasonal average and show that this surge is correlated with COVID case counts across states. By quantifying the number of excess ILI patients in March relative to previous years and comparing excess ILI to confirmed COVID case counts, we estimate the syndromic case detection rate of SARS-CoV-2 in the US to be less than 13%. If only 1/3 of patients infected with SARS-CoV-2 sought care, the ILI surge would correspond to more than 8.7 million new SARS-CoV-2 infections across the US during the three week period from March 8 to March 28. Combining excess ILI counts with the date of onset of community transmission in the US, we also show that the early epidemic in the US was unlikely to be doubling slower than every 4 days. Together these results suggest a conceptual model for the COVID epidemic in the US in which rapid spread across the US are combined with a large population of infected patients with presumably mild-to-moderate clinical symptoms. We emphasize the importance of testing these findings with seroprevalence data, and discuss the broader potential to use syndromic time series for early detection and understanding of emerging infectious diseases.

## 1 Introduction

The ongoing SARS-CoV-2 pandemic continues to cause substantial morbidity and mortality around the world [1, 2]. Regional preparation for the pandemic requires forecasting the growth rate of the epidemic, the timing of the epidemic peak, the demand for hospital resources, and the degree to which current policies may curtail the epidemic, all of which benefit from accurate estimates of the true prevalence of the virus within a population [3]. Confirmed cases are thought to be underestimates of true prevalence due to some unknown combination of patients not reporting for testing, testing not being conducted, and false-negative test results. Estimating the true prevalence informs the scale of upcoming hospital, ICU and ventilator surges, the proportion of individuals who are susceptible to contracting the disease, and estimates of key epidemiological parameters such as the epidemic growth rate and the fraction of infections which are sub-clinical.

The current literature suggests that the predominant symptoms associated with COVID are fever, cough and sore-throat; that is, patients often present with an influenza-like illness (ILI) yet test negative for influenza [4, 5]. With many COVID patients having a similar presentation as patients with influenza, existing surveillance networks in place for tracking influenza could be used to help track COVID.

Here, we quantify background levels of non-influenza ILI over the past 10 years and identify a recent surge of non-influenza ILI starting the first week of March, 2020. This surge of excess ILI correlates with known patterns of SARS-CoV-2 spread across states within the US, suggesting the surge is unlikely to be due to other endemic respiratory pathogens, yet is orders of magnitude larger than the number of confirmed COVID cases reported. Together this suggests that the true prevalence of SARS-CoV-2 within the US is much larger than currently appreciated and that the syndromic case detection rate is likely less than 13%, corresponding to more than 2.8 million new ILI cases due to SARS-CoV-2 and, assuming influenza-like clinical rates, more than 8.7 million total SARS-CoV-2 cases. Our analysis provides empirical corroboration of previous hypotheses of substantial undocumented cases yet places the estimated undocumented case rate higher than prior reports [6]. The SARS-CoV-2 prevalence estimates obtained from the ILI surge are consistent with an epidemic doubling time of less than 4 days. A sub-4 day doubling time is substantially faster than many prior reports [7, 8] yet is consistent with the 3-day doubling time of observed deaths due to COVID within the US. Our findings support a conceptual model for COVID spread in the US in which more rapid spread than previously reported is coupled with a larger undiagnosed population to give rise to currently observed trends. Finally, we find that the ILI surge peaks the week starting March 15, and we discuss the potential explanations for this phenomenon.

## 2 Results

### 2.1 Influenza like illness surge

We identified excess ILI cases by first subtracting cases due to influenza and then subtracting the seasonal signal of non-influenza ILI (Figure 1). Many states, including Washington, New York, Oregon, Pennsylvania, Maryland, Colorado, New Jersey, and Louisiana, have had a recent surge in number of non-influenza ILI cases far in excess of seasonal norms. For example, in the second week of March, 2020, Oregon saw 50% higher non-influenza ILI than it had ever seen since the inception of the ILINet surveillance system within the US. We find that with 95% probability, approximately 4% of all outpatient visits in Oregon during this time were for ILI that could not be explained by either influenza or the normal seasonal variation of respiratory pathogens. We find that as the seasonal surge of endemic non-influenza respiratory pathogens declines, this excess ILI correlates more strongly with state-level patterns of newly confirmed COVID cases suggesting that this surge is a reflection of ILI due to SARS-CoV-2 (Pearson *ρ* > 0.35 and *p* < 0.05 for the last three weeks; Figure S1A). Notably, we find that the ILI surge appears to peak during the week starting on March 15 and subsequently decreases in numerous states the following week; notable exceptions are New York and New Jersey, two of the states that have been the hardest hit by the epidemic, which have not started a decline by the week ending March 28.

**Figure 1:**
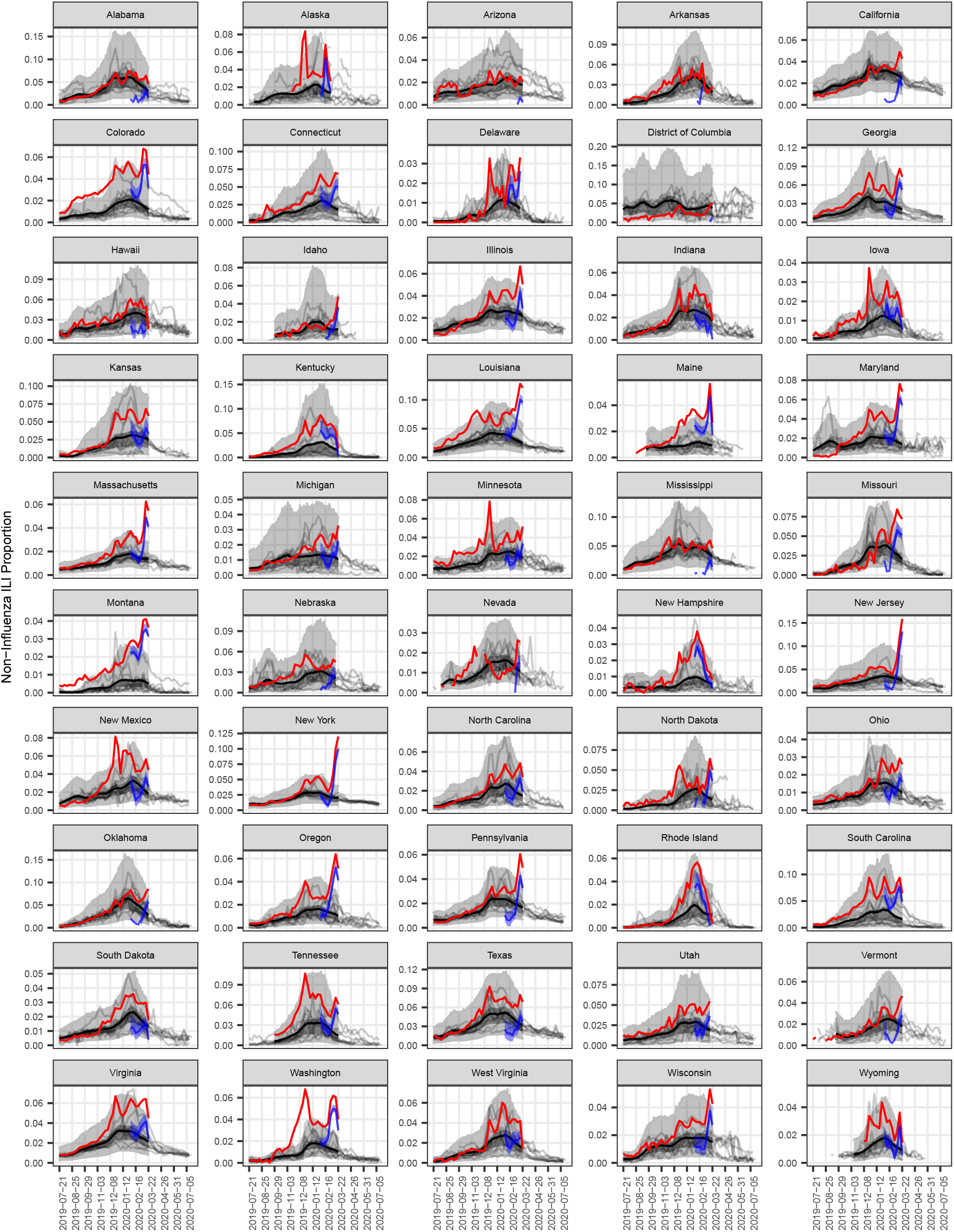
The excess non-influenza ILI is extracted from all non-influenza ILI by identifying the amount of non-influenza ILI in excess of seasonal norms (blue point and error bars represent the posterior median and 95% credible set for ILI not explained by non-COVID endemic respiratory pathogens). A binomial logistic-normal non-linear regression model was fit to non-influenza ILI data from 2010-2018 (grey lines). The model predicted the expected amount of non-influenza ILI in the 2019-2020 season (grey ribbons represent the 95% and 50% credible sets; the black line represents the posterior median). Observed non-influenza ILI beyond seasonal norms are shown as a blue line (posterior median) and blue ribbon (posterior 95% credible set). A number of regions are not represented due to insufficient laboratory influenza data to complete our analysis (see *Methods* for full details).

### 2.2 Investigating ILI Admission Rates

The magnitude of this ILI surge could be falsely elevated if patient behavior has recently changed leading to increasing detection of mild ILI. If the ILI surge reflected higher rates of detection of typically mild ILI, we would expect emergency department ILI rates would increase yet the proportion of those ILI cases admitted to the hospital would decrease. We were able to obtain data to evaluate this hypothesis from New York City’s influenza surveillance network [9]. In the month of March, the daily number of ILI visits to emergency departments across New York City increased while the proportion of those who went on to be admitted also increased by as much as 3-fold compared to the baseline rate prior to March (Figure S1). This suggests that patients are presenting less often for mild ILI, and such decrease in care-seeking behavior, if similar across the US, could be deflating the size of the ILI surge in later weeks of March.

### 2.3 Estimating Prevalence from the ILI Surge

To provide an estimate of the SARS-CoV-2 infections from the ILI Surge we assume that the population reported by sentinel providers is representative of their state each week. Additionally, we assume that the total number of patients needing medical care has not decreased since March 8th. Together these assumptions, as well as surveys describing the average number of patients seen by providers [10], the number of providers in each state [11], and the total number of outpatient visits per year [12, 13], allow us to estimate that, if outpatient clinics remained open during the COVID epidemics, we would expect that between March 8th and March 28th, there would have been approximately 2.8 million patient encounters with ILI due to COVID (95% credible set 2.6 million to 3.0 million; see *Methods* for full details).

Not all patients infected with SARS-CoV-2 will present to a health-care provider with ILI. While we cannot directly measure the rate of such sub-clinical cases, we suggest that a number of prior studies can be used to produce a lower-bound on this contribution. A recent study of passengers on the Diamond Princess cruise-ship accounted for right-censoring of patients sampled and estimated that 17.9% of patients infected with SARS-Cov-2 are asymptomatic for the course of their infection (95% credible set 15.5% to 20.2%). This estimate likely represents an underestimate given that the majority of passengers were >60 years old, a demographic thought to have a lower asymptomatic rate than younger individuals [14]. Beyond asymptomatic individuals, prior studies of ILI surveillance have suggested that only 20-50% of ILI cases seek medical care [15, 16]. To produce a lower-bound prevalence estimate we take the higher end of this range and assume that 40% of ILI cases present to a health-care provider. Together these additional contributions from sub-clinical cases produces a lower bound of 8.7 million SARS-CoV-2 infections between March 8th and March 28th (95% credible set 8.0 million to 9.4 million). Prevalence estimates for each state within this time-period are shown in Figure S4.

### 2.4 Syndromic Case Detection Rate

The rate at which SARS-CoV-2+ patients with ILI symptoms are identified as having COVID varies by state and over time (Figure S2). Our estimated syndromic case detection rates have been increasing over the month of March, which can be expected given increases in testing capacity across the US since the February 28 detection of community transmission in Washington State. For the week ending March 14, COVID cases in the states with the highest estimated syndromic case detection rate (Washington, Nevada, and Michigan) are only capturing approximately 1% of ILI surges in those states.In the last week ending on March 28, we estimate the detection rate across the US increased to be 12.5% (95% credible interval 9.5%-18.3%).

### 2.5 Epidemic Growth Rates and Clinical Rates

The true prevalence of SARS-CoV-2 is unknown at the time of this writing. However, if we assume the excess non-influenza ILI is almost entirely due to SARS-CoV-2, an assumption that becomes more valid as SARS-CoV-2 becomes more prevalent, we can use the excess non-influenza ILI to define bounds on the exponential growth rate of the US SARS-CoV-2 epidemic and understand the mutual dependence of exponential growth rates, the rate of subclinical infections, and the time between the onset of infectiousness and a patient reporting as ILI Figure 2. With a January 15 start date of the US epidemic [17], allowing early stochasticity from start-time to the onset of regular exponential growth, we find that it’s improbable to explain the ILI surge with an epidemic whose doubling time is longer than 4 days, as such slow growth scenarios fail to produce enough infected individuals to match the observed excess ILI.

**Figure 2:**
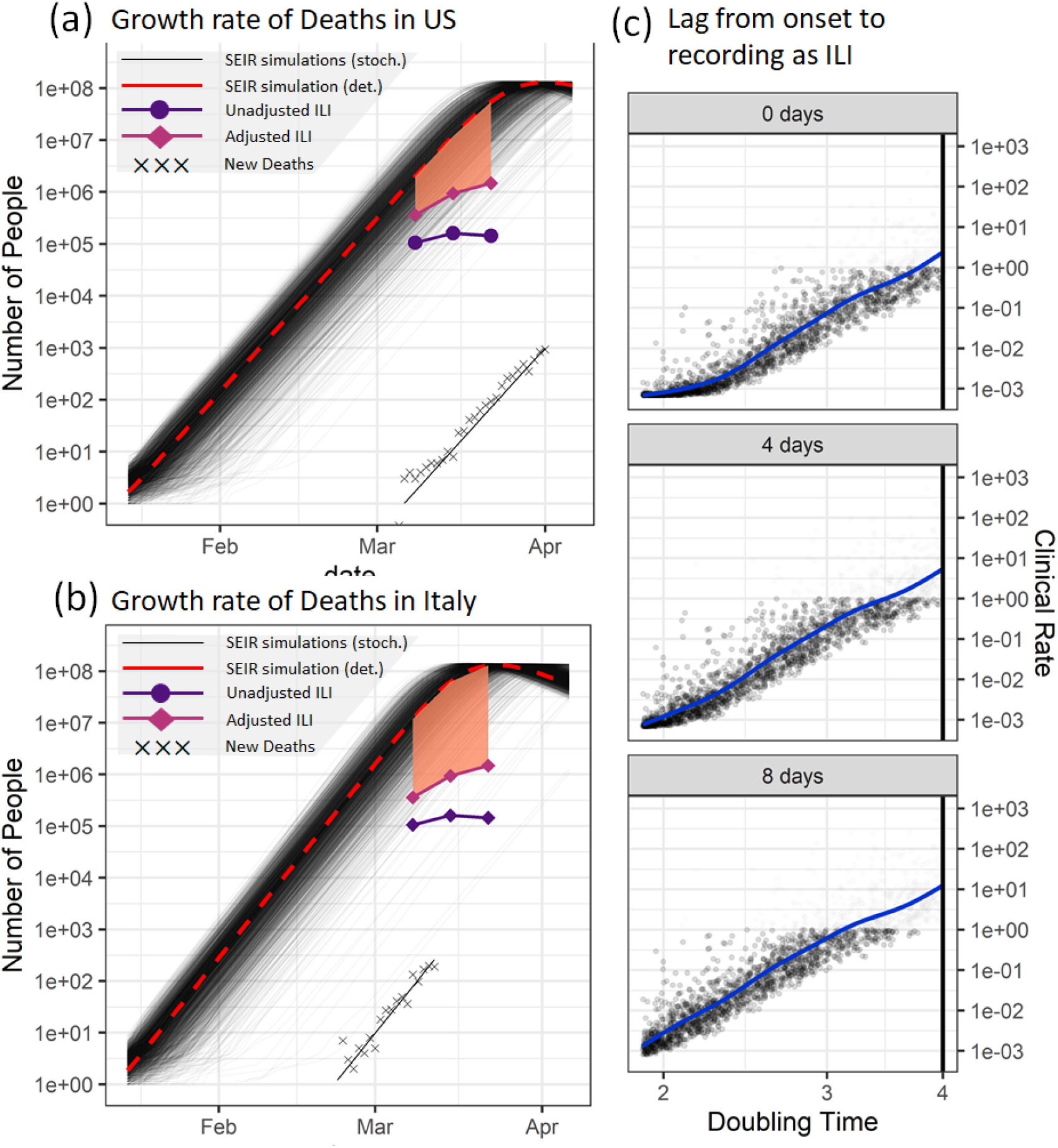
(a) The excess ILI estimated falls within the range of what one could expect from a US epidemic growing at the exponential growth rate defined by the growth of new deaths. Adjusting the ILI surge based on an estimated influenza-like subclinical rate, 17.9% asymptomatic infections and decreased care seeking in New York does not reconcile the difference in magnitude or slope between the ILI surge and the epidemic curve from US growth rates, suggesting additional forces are at play. The cause of the apparent deceleration in the ILI surge is hypothesized to be due to some combination of successful social distancing, faster decreases in care-seeking behavior than measured in New York, and/or other possibilities including faster growth and higher subclinical rates. (b) If the growth rate in the US is faster than US deaths suggest, such as a growth rate observed in Italy prior to the Italian lockdown (2.645 day doubling time), it could provide alternative explanations of the curvature of the excess ILI through a larger subclinical rate and epidemic curves near their peak at the time of the peak of the ILI surge. Serology or other measures of prevalence are needed to reconcile these alternative hypotheses. (c) More generally, the ILI surge forces a dependence between growth rate (doubling time), the clinical rate, and the lag between the onset of infectiousness and presentation to the doctor with ILI, where faster growth implies a slower clinical rate.

Across the entire US, new deaths due to COVID doubled every 3.01 days over the month of March (±0.001, p-value of test that doubling rate is less than 4 days approximately 0). An epidemic starting January 15th with doubling time equal to the doubling time of deaths in the US would imply an expected 10.4% clinical rate for the week of March 8 (the proportion of patients who have symptoms for which they would present to a health care provider) if the entirety of the first week of ILI surge is comprised of COVID patients and there was a 1-day lag from onset to presentation as ILI (Figure 3A). A four-day lag between the onset of infectiousness and presentation with ILI yields an average clinical rate of 23% among the 90.25% of simulations which could account for the ILI surge. However, the ILI surge did not grow at the same rate as deaths in the US, nor does an ILI surge adjusted to account for decreased care-seeking grow at the same rate as deaths, suggesting additional factors can be affecting the ILI surge, such as successful interventions, even faster decreases in care-seeking than observed in New York, or an early epidemic doubling faster than every 3 days.

Faster growth rates require lower clinical rates to explain the ILI surge. If the US epidemic prior to March 14 grew at the rate of deaths in Italy, doubling every 2.65 days, it could better match the curvature of the ILI surge by peaking around mid to late March, but would imply a clinical rate of 1.7% the second week of March (Figure 2B). If researchers produce alternative estimates of growth rates for the US epidemic, the ILI surge can be used to estimate bounds and ranges of possible clinical rates (Figure 3C). If the entirety of the ILI surge is attributable to COVID, it suggests a slowest-possible doubling time of 4 days for the US epidemic starting on January 15. Any evidence of significant secondary introductions, super-spreading, or rapid transmission events in early transmission chains will decrease these estimated clinical rates [18].

## 3 Discussion

We use outpatient ILI surveillance data from around the US to estimate the prevalence of SARS-CoV-2+. We find a clear, anomalous surge in ILI outpatients during the COVID epidemic that correlates with the progression of the epidemic across the US. The surge of non-influenza ILI outpatients is much larger than the number of confirmed case in each state, providing evidence of large numbers of symptomatic probable COVID cases that remain undetected. The slowest epidemic doubling time that could explain the ILI surge would be 4 days and this rate could only be achieved unusually fast early transmission or super-spreading events and a clinical rate near 100%. We measure the doubling time of deaths due to COVID with in the US to be 3.01 days and note that this is consistent with the bound imposed by the ILI surge. Together, the surge in ILI and analysis of doubling times suggest that SARS-CoV-2 has spread rapidly throughout the US since it’s January 15th start date and is likely accompanied by a large undiagnosed population of potential COVID outpatients with presumably milder distribution of clinical symptoms than estimated from prior studies of SARS-CoV-2+ inpatients.

Excess ILI appears to have peaked during the week starting on March 15th, leading the observed ILI dynamics to diverge from the overall epidemic dynamics implied by the growth rate of COVID deaths in the US. If the ILI dynamics were proportional to the epidemic curve then the two could be related with a constant subclinical rate. However, the changing ratio between the ILI surge and the epidemic curves parameterized by the growth rate of US deaths suggests additional mechanisms may be behind the ILI slowdown. First, a slowdown in ILI outpatient arrivals could be due to decrease in care-seeking where patients with mild ILI are less likely to present to the hospital as evident in emergency departments across New York City. Adjusting our ILI prevalence estimates based on the effect observed in New York City aligns ILI estimates more closely with predicted dynamics, yet the discrepancy remains. The remaining deviation could reflect more extreme changes in patient behaviors than those seen in New York City or successful interventions leading to lower transmission rates.

Our study has several limitations. First, the observed ILI surge may represent more than just SARS-CoV-2 infected patients. A second epidemic of a non-seasonal pathogen that presents with ILI could confound our estimates of ILI due to SARS-CoV-2. Alternatively, it is also possible that our use of ILI data has underestimated the prevalence of SARS-CoV-2 within the US. While early clinical reports focused on cough and fever as the dominant features of COVID [5], other reports have documented digestive symptoms as the complaint affecting up to half of patients with laboratory-confirmed COVID [19], and alternative presentations, including asymptomatic or unnoticeable infections, could result in ILI surges underestimating SARS-CoV-2 prevalence.

Additionally, our models have several limitations. First we assume that ILI prevalence within states can be scaled to case counts at the state level. This is based on the assumption that the average number of cases seen by sentinel providers in a given week is representative of the average number of patients seen by all providers within that state in a given week. Errors in this assumption would cause proportional errors in our estimated case counts and syndromic case detection rate. Second, our epidemic models are crude, US-wide SEIR models varying by growth rate alone and as such do not capture regional variation or intervention-induced changes in transmission. Our models were used to estimate growth rates from ILI for testing with COVID data and to estimate the mutual dependency of growth rate, the lag between the onset of infection and presentation to a doctor, and clinical rates; these models were not intended to be fine-grained forecasts for municipality hospital burden and other common goals for COVID models. Finer models with regional demographic, and case-severity compartments are needed to translate our range of estimated prevalence, growth rate, and clinical rates into actionable models for public health managers.

While an ILI surge tightly correlated with COVID case counts across the US strongly suggests that SARS-CoV-2 has potentially infected millions in the US, laboratory confirmation of our hypotheses are needed to test our findings and guide public health decisions. Our findings make the testable predictions that one would find relatively high rates of community seropositivity in states that have already seen an ILI surge and that seroprevalence of individuals infected in March is proportional to the size of the ILI surge. A study of ILI patients from mid-march who were never diagnosed with COVID could produce a focused test our predictions about the number and regional prevalence of undetected COVID cases presenting with ILI during that time. If seroprevalence estimates are consistent with our estimated prevalence from these ILI analyses, it would strongly suggest lower case severity rates for COVID than were assumed in late March and indicate the value of ILI and other public time-series of outpatient illness in facilitating early estimates of crucial epidemiological parameters for rapidly unfolding, novel pandemic diseases. Since not all novel pandemic diseases are expected to present with influenza-like symptoms, surveillance of other common presenting illnesses in the outpatient setting could provide a vital tool for rapidly understanding and responding to novel infectious diseases.

## 4 Methods

In what follows, let *i* index state *i* and let *t* index week *t* (with *t* = 0 referring to October 3, 2010; the start of ILINet surveilance).

### 4.1 Data Sources

Since 2010 the CDC has maintained ILINet for weekly influenza surveillance. Each week approximately 2,600 enrolled providers distributed throughout all 50 states as well as Puerto Rico, the District of Columbia and the US Virgin Islands, report the total number of patient encounters *n*_*it*_ and the total number of which met criteria for influenza-like illness (ILI – defined as a temperature 100F [37.8C] or greater, and a cough or sore-throat without a known cause of than influenza; *y*_*it*_) [20]. For scale, in the 2018-2019 season ILINet reported approximately 60 million outpatient visits. Coupled to these data are weekly state-level reports from clinical and public health labs detailing the number of patient samples tested for influenza 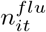 as well as the number of these samples which are positive for influenza 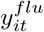. Therefore ILINet data can be thought of as a weekly state-level time-series representing the superimposed prevalence of various viruses which can cause ILI. ILINet data was obtained through the CDC FluView Interactive portal [21].

In addition to ILINet data, US State population data for the 2020 year was downloaded from https://worldpopulationreview.com/states/. The number of primary care providers in each state per 100,000 residents *b*_*i*_ was obtained from the United Health Foundation [11]. COVID confirmed case counts were obtained from The New York Times’ database maintained at https://github.com/nytimes/covid-19-data. This dataset contains the daily cumulative confirmed case count for COVID for each state *z*_*il*_ for day *l*. The dataset of deaths in Italy was downloaeded from https://github.com/pcm-dpc/COVID-19 on April 6, 2020.

### 4.2 Data Processing

Within the ILINet dataset, New York City and New York were summed into a combined New York variable representing both New York city and the surrounding state. Due to incomplete data in one or more of the data-sources described above the Virgin Islands, Puerto Rico, The Commonwealth of the Northern Mariana Islands, and Florida were excluded from subsequent analysis. In addition, daily cumulative confirmed COVID cases were converted to weekly counts of new cases by

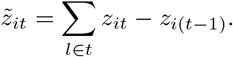

### 4.3 Extracting non-influenza ILI signal

To subtract influenza signal from *y*_*it*_ we assume that the population of patients with ILI within a state are the same population that are potentially tested for influenza. This assumption allows us to calculate the number of non-influenza ILI cases as

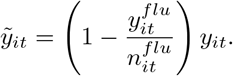

The resulting time-series 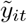 are shown in Figure S3.

### 4.4 Identifying ILI Surges

We identified ILI surges in 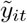 by training a model on 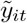 for all data prior to July 21, 2019. We then used this model to predict the prevalence of non-influenza ILI 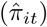 for dates after and including July 21, 2019. We calculated the ILI surge as the difference between the observed proportion of non-influenza ili 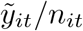 and 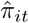.

More specifically, to account for variation in the number of reporting providers, we trained the following binomial logistic-normal model

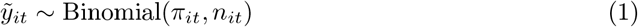

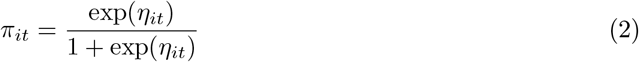

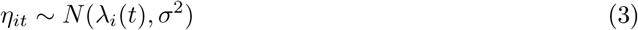

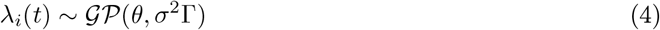

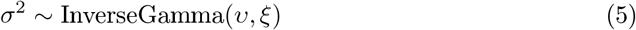

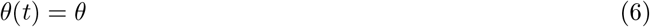

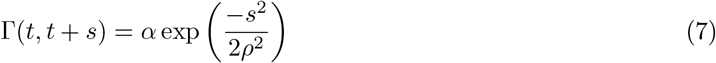

We made the following prior specifications: We set the bandwidth parameter for the squared exponential kernel as *ρ* = 3 representing a strong local correlation in time that died off sharply beyond 3 weeks, *α* = 1 representing a signal to noise ratio of of approximately 1, *υ* = 1 and *ξ* = 1 representing weak prior knowledge regarding the overall scale of variation in the the latent space. Finally, we set *θ* = −2.197 representing an off-season prevalance of 0.1% non-influenza ILI. Samples from the posterior predictive density 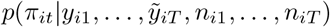 were collected using the function *basset* from the R package *stray* [22]; a total of 4000 such samples were collected in this analysis. We define the prevalence of non-influenza ILI in excess of normal seasonal variation as 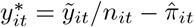.

To exclude variation attributable to unseasonably high rates of other ILI causing viruses (such as the outbreak of RSV in Washington state in November-December 2019) we only investigate 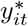 for weeks after March 7th 2020 as only these later weeks had high correlation to the COVID confirmed case rate (Figure S1).

### 4.5 Calculating scaling factors to relate ILINet data to COVID cases

As COVID new case counts 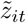 represent the number of confirmed cases in an entire state and ILINet data represents the number of cases seen by a select number of enrolled providers, we must estimate scaling factors *w*_*i*_ to enable comparison of ILINet data to confirmed case counts at the state level. Let 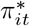 denote the probability that a patient with ILI in state *i* has COVID as estimated from ILINet data. Let *p*_*i*_ denote the population of state *i* and let *b*_*i*_ denote the number of primary care providers per 100,000 people in state *i*. We translate the inferred proportion of individuals with ILI due to COVID to the state level by considering the average number of patients seen across all providers in the state in a 5 day work-week. In addition, we add a discount factor *λ* = 0.55 to calibrate these estimates with prior reports regarding the total number of outpatient visits per year [13]. This yields our estimated number of COVID cases (excess ILI at the state level) as

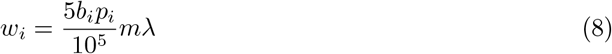

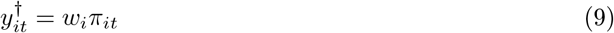

where *m* = 20.2 is the mean number of patients seen by physicians per day [10].

### 4.6 Accounting for sub-clinical infections

To account for the contribution of sub-clinical SARS-CoV-2 infections we use a recent analysis of cohort surveillance from the Diamond Princess [23]. Monte-Carlo simulations were used to propagate error from our uncertainty regarding potential asymptomatic infections affecting the clinical rate *δ*^*b*^ into our calculation of posteriors for epidemic trajectories. To match posterior estimates from Mizumoto et al.[23], we use quantile matching to parameterize *δ*^*c*^∼ Beta(*α, β*) to achieve a mean of .179 and a 95% probability set of (.155, .202). In addition, we take *δ*^*c*^ = 0.4 as a conservative estimate of the proportion of patients with ILI who would seek medical care in a typical year [15, 16]. To account for these sub-clinical contributions we use adjusted scaling factors

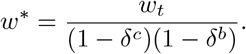

### 4.7 Estimating syndromic case detection rates

Assuming that the majority of SARS-CoV-2 testing within the US has been directed by patient symptoms[24], the pool of newly diagnosed SARS-CoV-2+ patients is a subset of the pool of SARS-CoV-2+ patients who are identified as having ILI. Therefore, we calculate the probability that a SARS-CoV-2+ patient with ILI who seeks medical care will be identified as having SARS-CoV-2 as 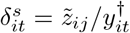 (Figure S2).

### 4.8 Growth Rate estimation

Growth rates were estimated for the US and Italy by poisson generalized linear models predicting new deaths with date. Data on COVID deaths in the US were obtained from https://github.com/nytimes/covid-19-data on April 6, 2020 and all deaths from March 5, 2020 to April 1, 2020, were summed by date. Initially, April 2-5 were included but were found to have extremely high leverage and were hence excluded from our analysis. Data on COVID deaths in Italy were obtained from https://github.com/pcm-dpc/COVID-19. The same procedure was applied, focusing on deaths from February 24 until March 12. The slope from poisson regression was used as the estimated exponential growth rate, yielding a US growth rate *r*_*US*_ = 0.23 or a 3.01 day doubling time and *r*_*IT*_ = 0.26 or a 2.65 day doubling time.

### 4.9 Epidemic simulations and clinical rates

SEIR models,

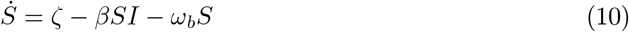

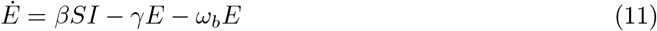

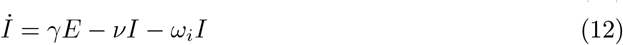

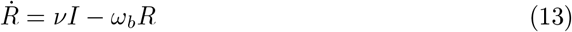

were parameterized for the US to a timescale of units days by setting *ζ* = 3.23 × 10^−5^ corresponding to a crude birth rate of 11.8 per 1000 per year, a baseline mortality rate *ω*_*b*_ = 2.38 × 10^−7^ corresponding to 8.685 per 1000 per year, an infectious mortality rate *ω*_*i*_ = 4.96 × 10^−4^, corresponding to a infection fatality rate of 0.5% required to fit US deaths under a 20-day lag from osnet to death. Further, we used an incubation period *γ*^−1^ of 3 days, infectious period *ν*^−1^ of 10 days, and *β* parameterized to ensure *I*(*t*) grew with a specified exponential growth rate early in the epidemic. A total of 2,000 simulations were run for each of the two growth rates (US and Italy) analyzed. Growth rates were drawn at random with *r*_*US*_ *N* (*r*_*US*_, 0.1) and *r*_*IT*_ *N* (*r*_*IT*_, 0.1). To illustrate the mutual dependence between estimates of growth rate, clinical rate, and the lag between the onset of infectiousness to presentation to a doctor with ILI, 2,000 simulations with uniform growth rates in the interval [0.173,0.365] corresponding to a range of doubling times between 1.9 days and 4 days.

Each simulation was initialized with (*S, E, I, R, t*) = (3.27 × 10^8^, 0, 1, 0, 0) where time 0 was January 15. To simulate the stochastic time it took from the first case to the onset of regular exponential growth, a Gillespie algorithm was used from the initial conditions until either *t* = 50 (March 5, 2020) or *E*(*t*) + *I*(*t*) = 100. The initial Gillespie algorithm was implemented on the assumption that a large amount of variation in the epidemic trajectory stems from uncertainty in trajectory of early transmission chains. The output from Gillespie simulations was input as an initial value into the system of differential equations and integrated until the August 5, 2020. The number of infected individuals on a given day was the last observed *I*(*t*) for that day, and a weekly pool of infected patients was computed by a moving sum over the number of infected individuals every day for the past week, 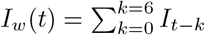.

Defining 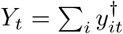 as the national excess ILI, the clinical rate implied by a given simulation was estimated as

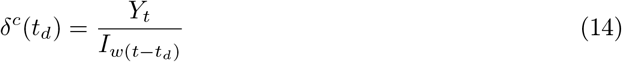

for a given time delay *t*_*d*_ it takes from the onset of infectiousness to a patient reporting to the doctor with ILI.

## Data Availability

All analysis files and datasets, save large US SEIR simulations, available on GitHub at https://github.com/jsilve24/ili_surge

## 4.10 Code Availability

All code and data required to reproduce our results is publicly available at https://github.com/jsilve24/ili_surge.

## 5 Acknowledgements

We thank Rachel Silverman, Raina Plowright, Colin Parrish, Dan Rosenheck, and Seth Stephens-Davidowitz for their manuscript comments. JDS was supported in part 340 by the Duke University Medical Scientist Training Program (GM007171).

---

**Figure S1:**
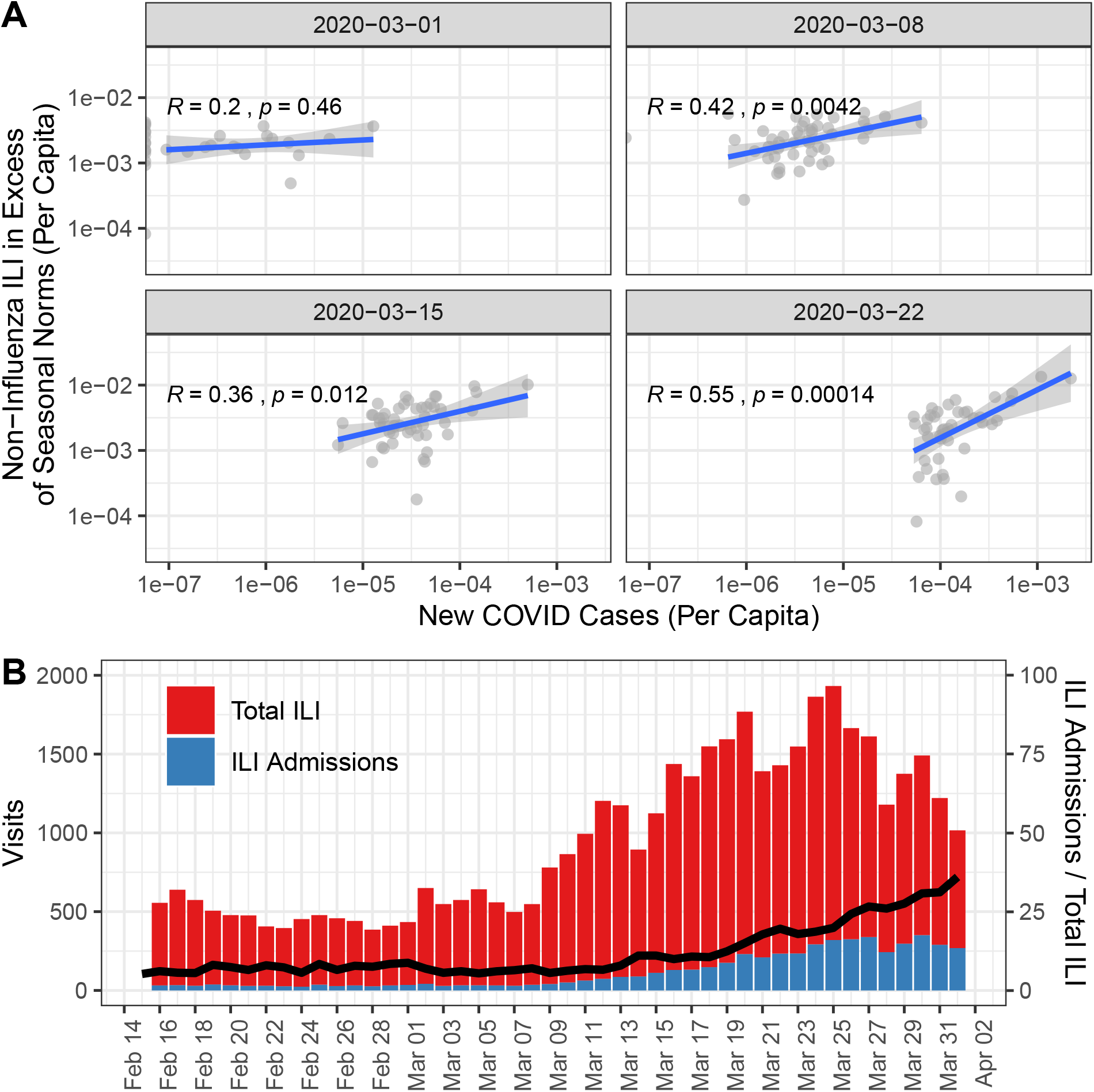
(A) Excess ILI correlates strongly with patterns of newly confirmed COVID cases. This correlation is strongest for the last three weeks of data, when other seasonal respiratory pathogens are at their lowest. (B) Of all ED visits for ILI in New York City, the proportion (black line) of those severe enough to warrant admission to the hospital has increased in the past month.

**Figure S2:**
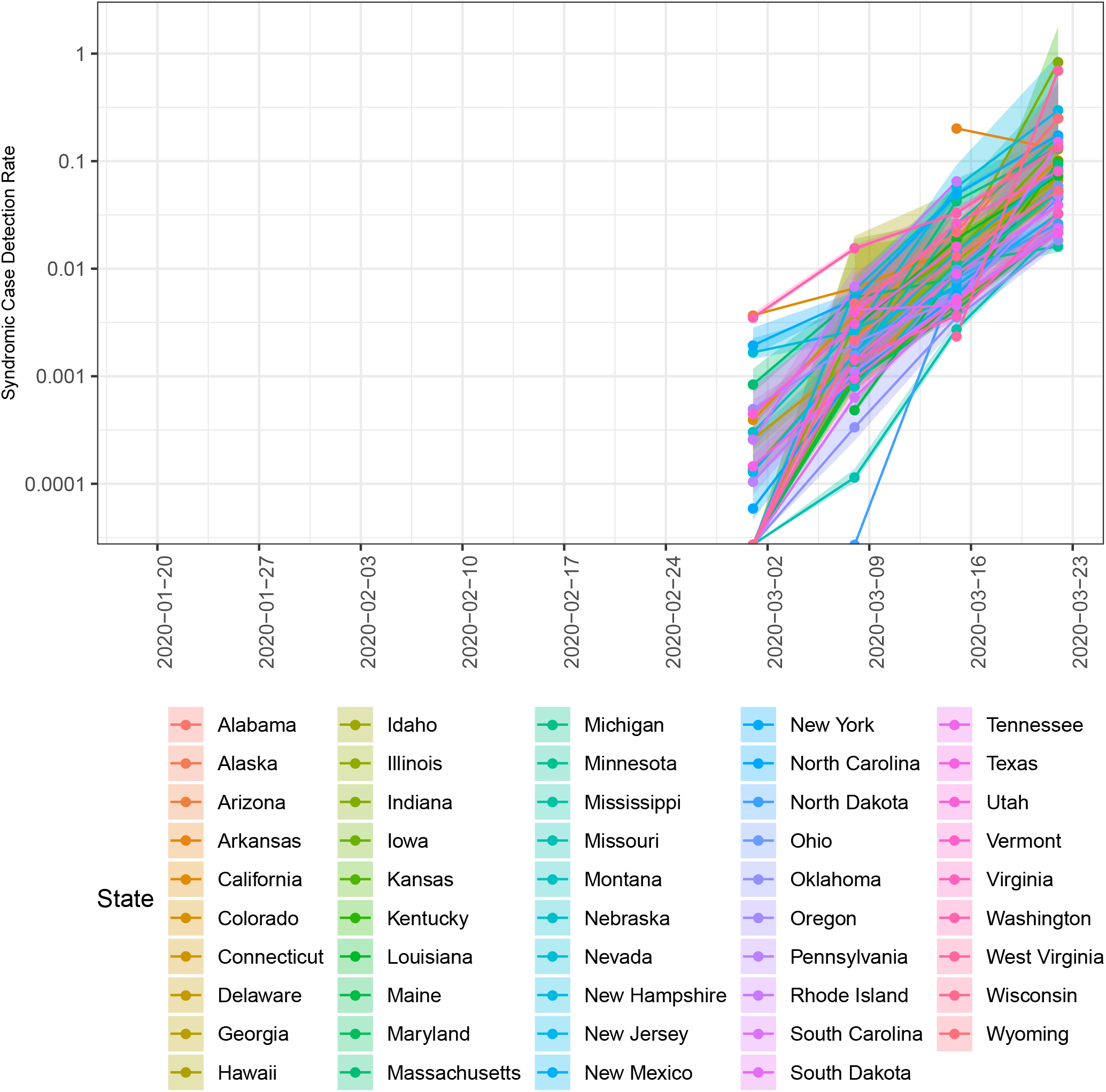
Since March 1, 2020, the case-detection of symptomatic COVID patients has increased by a factor of ≈ 100. This likely represents increased awareness of community transmission within the US combined with increased availability of testing. Still, the syndromic case detection rate remains below 1% for most states with many states with detection rates closer to 0.1%.

**Figure S3:**
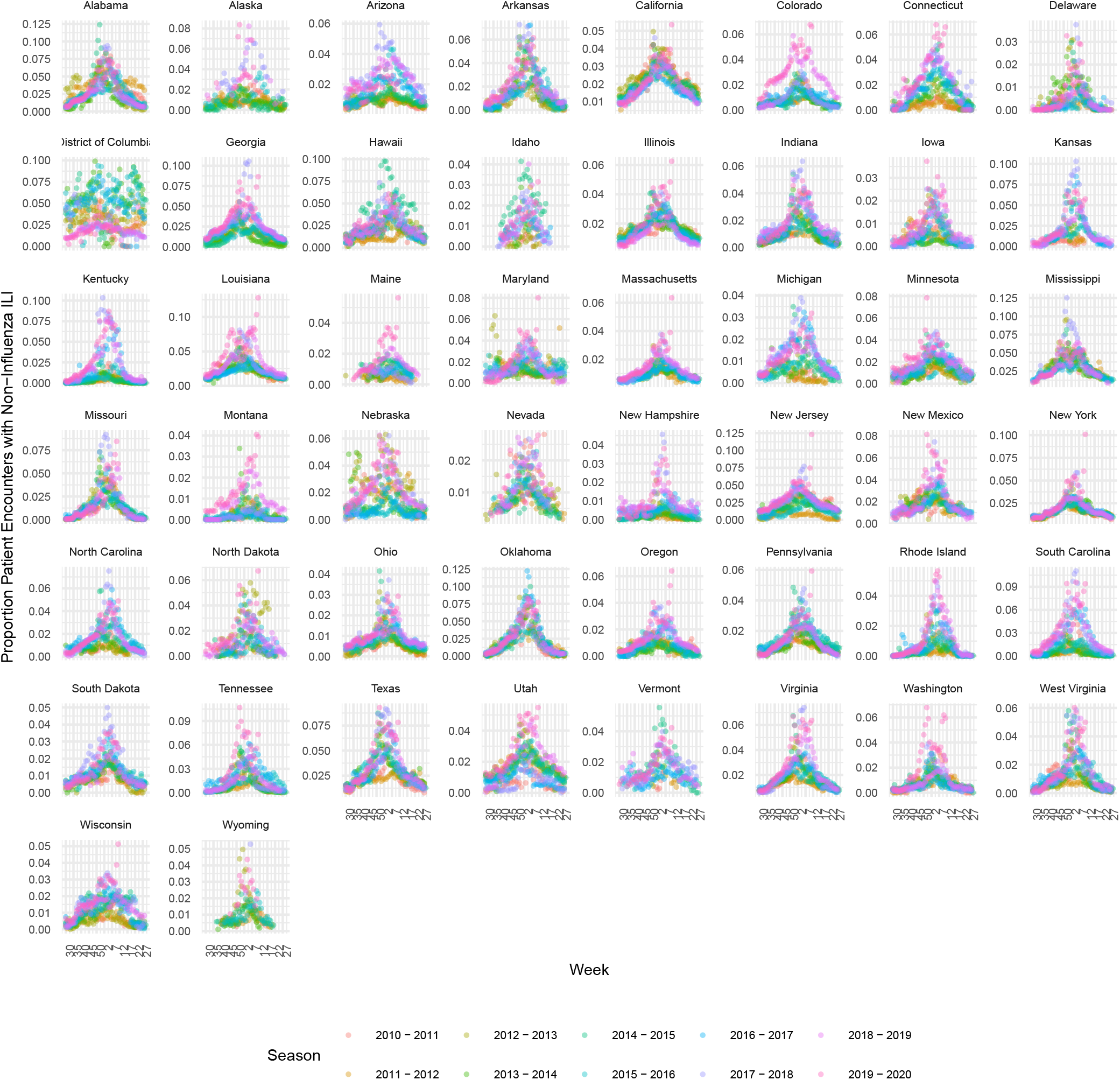
Once the signal attributable to influenza is extracted, the proportion of Patient encounters in which patient had non-influenza ILI (*y*∼_*it*_*/n*_*it*_) displays strong seasonal trends. The most notable deviations from these trends occur around Febuary to March of the 2019-2020 flu season and align with the onset of the COVID epididemic within the US.

**Figure S4:**
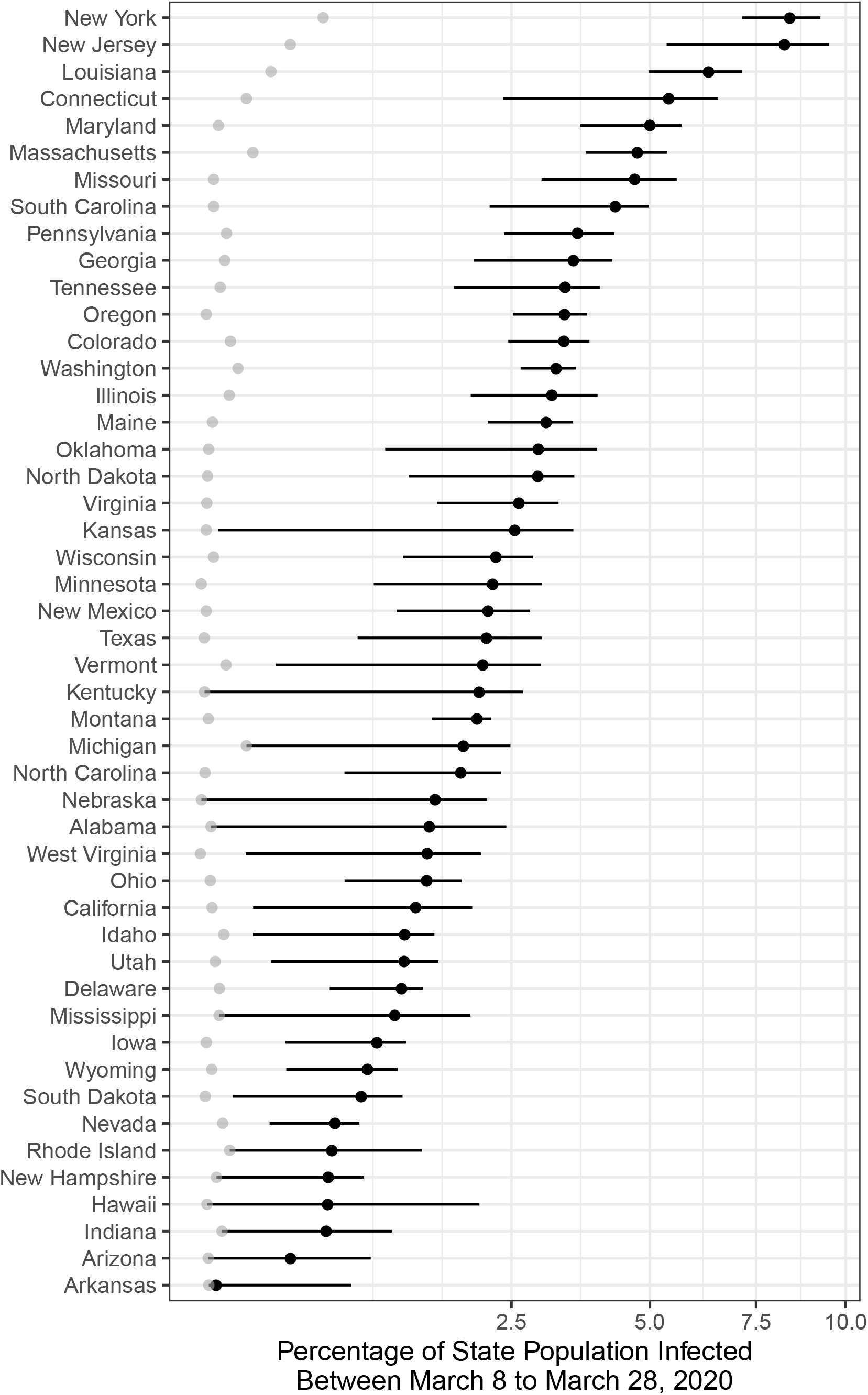
Prevalence of SARS-CoV-2 infections between March 8 and March 28, 2020 (posterior median and 95% credible set, black). Corresponding estimates based on confirmed case counts are shown in Grey.

